# Double Trouble - The prevalence of concomitant traumatic brain injury in individuals with spinal cord injury and its impact on functional outcomes: a systematic review

**DOI:** 10.1101/2025.10.07.25337129

**Authors:** Keziah Skein, Rebecca George, Ryan L. O’Hare Doig, Frances Corrigan, Anna V Leonard

**Affiliations:** Translational Neuropathology Laboratory, School of Biomedicine, Faculty of Health and Medical Sciences, Adelaide University; Adelaide Medical School, Adelaide University, Adelaide, SA 5005, Australia; Neil Sachse Centre for Spinal Cord Research, Lifelong Health Theme, South Australian Health and Medical Research Institute, Adelaide, SA 5000, Australia

**Author notes:** These authors contributed equally to this work (Frances Corrigan, Anna V Leonard).

**Keywords:** Traumatic brain injury, spinal cord injury, incidence, motor, cognition, systematic review

## Abstract

**Study design:** Systematic review

**Objectives:** To examine the prevalence, diagnostic challenges, and functional impact of concomitant traumatic brain injury (TBI) in individuals with traumatic spinal cord injury (SCI).

**Methods:** PubMed, Embase and Scopus databases were searched with a search strategy containing key search terms for TBI, SCI and concomitant injury. Original research articles reporting on prevalence and/or functional outcomes following a TBI at the time of SCI in adult populations were included.

**Results:** Forty studies met the inclusion criteria, with 32 reporting prevalence and 18 information on functional outcomes. Reported prevalence rates of concomitant TBI varied widely (10–75%) across studies, largely due to inconsistent diagnostic criteria, retrospective data collection, and reliance on incomplete medical records or ICD coding. The identification of mild TBI (mTBI) was particularly problematic, with differing diagnostic criteria employed.. Moderate–severe TBI at the time of SCI significantly increased in-hospital mortality and complications like pneumonia, sepsis, but had minimal effects on rehabilitation trajectory. Functional outcomes, particularly motor and sensory recovery, were generally unaffected by concomitant injury, though subtle cognitive deficits were observed in moderate to severe TBI cases during rehabilitation. Few studies examined outcomes beyond one year post injury.

**Conclusion:** Overall, current evidence suggests that concomitant TBI is common in people presenting with an SCI, but its long term functional and cognitive impact remains underexplored. Future research should employ standardised diagnostic criteria, prospective data collection, and long term follow up to clarify the role of concomitant TBI not only in the acute recovery phase, but also chronically.

## 1. Introduction

Spinal cord injury (SCI) is a global issue with national incidence reported at ∼900 000 cases/year^1^, leading to ∼21 million people living with an SCI^2^. Following SCI, neuronal loss at the injury site results in either a complete or partial motor control deficit below the level of injury, as well as a loss of sensory input to the brain from areas below the injury level^3–5^. Additionally, damage to neurons at the injury site disrupts the entire autonomic nervous system^3,6^, manifesting predominantly as impairments in thermoregulation, bladder control, and bowel function^3,5^.

Beyond the symptoms associated with direct disruption of motor and sensory pathways SCI is also linked to an impairment in cognitive function, the development of neuropathic pain, and mood changes. Cognitive function impairment has been observed in both clinical and pre-clinical research^7^, with people with an SCI exhibiting significantly lower cognitive performance compared to healthy controls when assessed for attention, memory, executive function, visuoconstructive ability, and language^8^. This impairment is seen shortly after the SCI and persists for at least 1 year following the injury^9,10^. Additionally, SCI causes diminished information processing capabilities^11^, with people reporting significantly elevated chronic fatigue levels after engaging in cognitive tasks after experiencing an SCI^12^. SCI is also associated with the development of neuropathic pain in 35-41%^13,14^ of people, resulting in the experience of severe sharp, shooting, and burning sensations in response to stimuli that previously was either innocuous (allodynia) or minorly noxious (hyperalgesia)^15^. This in part relates to alterations in the pain signalling pathways, including not just the spinal cord, but also regions of the brain responsible for pain regulation^15–17^. Indeed, SCI is associated with increased incidence of other neurological conditions including depression and anxiety^12,18^, with the incidence increasing significantly over the first six months post-rehabilitation discharge^8^.

One confounding factor that may play a role in recovery following an SCI, is the presence of a concomitant traumatic brain injury (TBI). The biomechanics of the types of insults that cause SCI are like those related to TBI, like falls and motor vehicle accidents (MVAs). A concomitant TBI may exacerbate SCI associated neurological deficits, with TBI also leading to severity-dependent motor symptoms like paresis, ataxia, or postural instability^19^, cognitive symptoms like information processing^20^, recognition memory^21^, episodic memory^22^, or verbal learning and memory^23^, and the development of neuropathic pain^24^. This may be driven by a concomitant TBI influencing the pathological response to the SCI. Both TBI and SCI are characterised by a primary insult, the direct physical injury, followed by a prolonged secondary injury phase^25^. The secondary injury phase involves a robust neuroinflammatory response, characterised by activation of resident cells, migration and recruitment of leukocytes and the release of inflammatory mediators^26^. Inflammation is not localised to the site of injury with release of mediators including TNFα, IL-10, CCL2 and IL-8 within the CSF and serum^27–29^. Given the widespread release, a local inflammatory response within the brain may influence that within the spinal cord and vice versa. In support of this, mild TBI has been shown to induce inflammatory changes with neuronal loss within the remote spinal cord^25,30^ and thoracic SCI was associated with microglial changes chronically throughout the brain with neuronal loss^17,31^, and the later development of cognitive deficits^31^.

However, it is difficult to determine the impact of a TBI on SCI recovery, given that reports of the incidence of concomitant TBI vary widely from 7.4-74.2%^32^, with little known about what drives this disparity across different studies. Additionally, up to 60% of TBI diagnoses may be missed in those presenting with an SCI to emergency care^33,34^. As such this systematic review aimed to examine prevalence of concomitant SCI and TBI, factors influencing the differences in reported prevalence between studies, and whether concomitant injury influenced functional recovery post-SCI. This builds upon previous reviews^35,36^, by integrating evaluation of incidence of a double diagnosis of SCI with TBI (DDx), methods for screening of co-occurring TBI and potential impacts of DDx on functional outcomes.

## 2. Methods

### 2.1 Search Strategy

The Preferred Reporting Items for Systemic Reviews and Meta-Analysis (PRISMA) guidelines were used. A comprehensive search was initially performed in November 2022 and updated in April 2023 and October 2024, using the electronic databases Medline via PubMed, EMBASE, and Scopus to identify relevant publications. The search strategy was developed in consultation with a health and medical sciences librarian and used the terms “Traumatic brain injury”, “Spinal cord injury” and “Concomitant” and related synonyms (Supplementary material S1). Additional searching of reference lists was also performed to identify further relevant studies.

### 2.2 Selection Criteria

Following the database search, all citations were collated and uploaded into EndNote (EndNote™ X9) and then imported into Covidence systematic review software (Veritas Health Innovation, Melbourne, Australia). Duplicates were identified, removed and cross checked.

The title and abstracts of the resulting publications were screened and included if they included reference to both TBI and SCI in a clinical population. For articles that passed this preliminary assessment, the full text article was retried and screened for eligibility against the inclusion and exclusion criteria as assessed by two independent reviewers, with conflicts resolved by a third reviewer. A flow chart with reasons for exclusion of studies is displayed in **Figure 1**.

**Fig 1:** PRIMSA flow diagram outlining the article selection and screening process and subsequent data management.

#### 2.21: Inclusion criteria

The following inclusion criteria were utilised:

i. An original research article published in English
ii. Investigated a predominantly adult population, 16 years or older
iii. TBI occurred at the time of the SCI
iv. Provided information on the incidence of DDx and/or information on the influence of DDx on functional recovery
v. Published in the last 40 years

#### 2.2.2 Exclusion criteria

i. Did not state when the TBI occurred relative to the SCI
ii. Did not include both a DDx and a SCI group
iii. Study did not differentiate between spinal and spinal cord injury
iv. Studies only focused on a subpopulation of those with an SCI (ie cervical injury only)
v. SCI population was a mix of non-traumatic and traumatic causes
vi. Studies were single case reports/expert opinions
vii. Studies were review articles or conference abstracts.
viii. Full-text unavailable
ix. Pre-clinical study only

### 2.3 Data Extraction

A data collection spreadsheet was then developed to extract the relevant information from the included articles. Data extracted included study characteristics (author, year of publication, study design, location), participant characteristics (sample size, age, sex, time-point measured post-injury, mechanism of injury, severity of SCI, severity of TBI, criteria used to diagnose TBI) and functional outcome information where available (test/s used, time-point, time in hospital, time in rehabilitation). Given the diversity of functional outcomes examined, these were classified as cognitive, autonomic, motor, psychological or other where they did not fit in any of these categories.

### 2.4 Assessment of methodological quality

Articles included in the study were assessed for methodological quality by one reviewer (FC), with confirmation provided by a second reviewer (RG), by using an adapted version of the National Institutes of Health quality assessment tool (Supplementary material S2)^37^. A study was deemed to be of high quality if it scored greater than 70%, moderate quality with scores between 50 and 70% and low quality when scoring below 50.

## 3. Results

### 3.1 Search results

Electronic searches of PubMed, Embase and Scopus databases produced a total of 1067 titles. 454 duplicates were removed leaving 613 titles. On reviewing titles and abstracts, a further 395 records were excluded, with 218 articles sought for retrieval. 18, were unable to be obtained, with review of the remaining articles identifying a further 6 articles via citation analysis., leaving 206 articles to be assessed for eligibility. Full-text review excluded a further 166 articles, which did not meet the inclusion criteria leaving 40 articles meeting the inclusion criteria. Details of this process are described in the PRISMA diagram (Figure 1).

### 3.2 Characteristics of included studies

Of the relevant studies 32 provided a prevalence estimate of the incidence of DDx in people with an SCI and 18 information on functional outcomes. The majority (26/40) had the primary aim of investigating DDx, while in 14 papers information on DDx was presented as one of several outcomes reported. The majority were cross-sectional (n=20) or cohort designs (n=17), with a small number of case control studies (n=3). There was an even mix of prospective (n=16) and retrospective (n=21) population recruitment, with 3 studies utilising both. Most of the studies were conducted in North America, in the US (n=17) and Canada (n=7), with other studies were conducted in Europe (n=10), Asia (n=2), Oceania (n=3) and South America (n=1). Sample sizes were generally small, with six studies with less than 50 participants, six with 50-100 participants, seven with 101-500 participants, five with 501-1000, four with 1001-2000 and only three with more than 10,000 participants.

### 3.3 Methodological quality

A summary of the methodological quality assessment for the included articles is provided in Fig. 2 and Supplementary Table 3. Fifteen (37.5%) studies were of high quality, ten (25%) of moderate quality and 15 (37.5%) of low quality, with sample size justification, consideration of the severity of TBI, identification of TBI and statistical analyses the main domains of concern.

**Fig 2:** Assessment of methodological quality presented as the number of studies exhibiting each attribute.

### 3.4 Diagnostic criteria

As identified within the methodological analysis, studies varied in how the presence of a concomitant TBI was identified, with the majority relying on retrospective analyses, utilising predominantly hospital medical records (n=26), but also national trauma data banks (n=4), clinical study data (n=1) or rehabilitation admission records (n=1), with one study not specifying the source of information^38^ (Table 1). Four of these studies also specified the inclusion of emergency medical service, surgical and acute rehabilitation records alongside their hospital medical record review to increase the likelihood of finding relevant information ^33,39–41^. Of the studies that utilised a prospective TBI diagnosis, in two studies this occurred at admission to hospital^42,43^, and in five at admission to rehabilitation^9,10,44–46^. A further four studies used both retrospective record review and prospective clinical examination encompassing neuroimaging^34^, the step 2 TBI-4 Self Report interview^47^ or neuropsychological testing^46,48^.

**Table 1:** Diagnostic criteria utlised to identify the presence of concomitant TBI. Data is presented from lowest to highest incidence in the prevalence studies, with studies grouped as an incidence <30% (green) 30-50% (blue) or >50% (orange).

|  | Diagnostic criteria |  |  |  |  |  |  | Determination | Data source |
| --- | --- | --- | --- | --- | --- | --- | --- | --- | --- |
|  | PTA | LOC | GCS | Neuroimaging | Symptoms/signs | Billing codes | Other |  |  |
| Prevalence/Incidence studies |  |  |  |  |  |  |  |  |  |
| Varma, 2010 |  |  |  |  |  | ICD-9 (not specified) |  |  | Retrospective, hospital medical records |
| Ramamurthy, 2017 | Present | Mild < 5mins<br>Mod:5-24h<br>Sev:>24h | Mild: 14-15<br>Mod: 9-14<br>Severe: 3-8 |  | Mild: - focal neurological deficits<br>Mod/Sev: +ve Focal neurological deficits |  |  | Only one criterion required for diagnosis | Retrospective, hospital medical records |
| Azad, 2024 |  |  |  |  |  | ICD-9, 10 (700, 801, 803, 804, 850, 851, 852, 853, 854, S02.0, S02.1, S02.91, S04.02, S04.03, S04.04, S06, S07.1) |  |  | Retrospective, National Trauma Data Bank |
| Furlan, 2023 |  |  | <15 |  |  |  |  |  | Retrospective, NASCIS 3 study |
| Melo Nelo, 2014 |  |  | Mild: 13-15<br>Mod:9-12<br>Sev:3-8 |  |  |  |  |  | Prospective, Hospital admission |
| Dvorak, 2023 |  |  |  |  |  | ICD-10 (not specified) |  |  | Retrospective, Spinal Cord/Trauma registry |
| Strubreither, 1997 |  |  |  |  | Not specified |  |  |  | Retrospective rehab admission records |
| Tachino, 2024 |  |  | Mild: 13-15<br>Mod:9-12<br>Sev:3-8 |  |  |  |  |  | Retrospective, Japan trauma bank |
| Creasey, 2015 |  |  |  |  |  | ICD-9 (not specified) | Reference to 'TBI', 'brain injury', 'brain trauma', 'head injury', 'head trauma' or 'LOC' |  | Retrospective, hospital medical records |
| Saboe 1990 |  |  |  |  |  |  |  |  | Retrospective, hospital medical records |
| Yang, 2023 | Mild <24h<br>Mod 1-7d<br>Sev >7d |  | Mild: 13-15<br>Mod:9-12<br>Sev:3-8 | Mod:sev +ve | Mild: MoCA>18<br>Mod: MoCA 14-18<br>Sev MoCA <14 |  |  | Either PTA or LOC | Retrospective, hospital medical records |
| Brougham, 2011 |  |  |  |  |  | ICD-9 (not specified) |  |  | Retrospective, hospital medical records |
| *Wei, 2008* |  |  |  | MRI +ve |  |  |  |  | Prospective, rehab centre |
| *Bombardier, 2016* | Present |  | Mild:13-15<br>Mod:9-12<br>Sev:3-8 | CT+ve | Mental status change |  | Resident/attending diagnosis | GCS<15 = always TBI<br>If GCS=15 need +CT or PTA | Retrospective, hospital medical records |
| *Mansfield, 2014* |  |  |  |  |  | ICD-9: 800.0-804.99,<br>850.0-854.19, 959.0 |  |  | Retrospective, US trauma bank |
| *Mirzaeva, 2022* |  |  |  |  |  | ICD-10, code S06 |  |  | Retrospective, hospital medical records |
| *Budisin, 2016* | Present | Present | Present | Positive |  |  |  | Require 2 of PTA, LOC and GCS of 14 or below OR positive neuroimaging. | Retrospective, hospital medical records<br>Prospective: Neuroimaging |
| *Davidoff, 1985* | Present | Present |  |  |  |  |  | Only one of the criteria needed for diagnosis | Prospective, interview on admission to rehab |
| *Nott, 2014* |  | Present | <15 | Abnormal | Positive |  |  | Only one of the criteria needed for diagnosis | Retrospective, hospital medical records |
| *Mirzaeva, 2019* |  |  |  |  |  |  |  |  | Retrospective, hospital records |
| *Mirzaeva, 2020* |  |  |  |  |  |  |  |  | Retrospective, hospital medical records |
| *Garlanger, 2018* | Mild: <24h<br>Mod/Sev: >24h | Mild <30 mins<br>Mod/Sev: >30 mins | Mild: 13-15<br>Mod/Sev: <13 | Evidence of Skull fracture, intracranial haematoma, contusion |  |  |  | Only one of the criteria needed for diagnosis | Retrospective, hospital records, neuroimaging, neuropsychology notes and rehabilitation notes |
| *Gordan, 2012* |  | Present |  |  |  | ICD-9: 800–801.99, 803–804.99, 850–854.19, 873.0–873.1, 873.8–873.9, 905.0, 907.0, 959.01 | Reported brain injury complication e.g SAH contusion, concussion | Only one of the criteria needed for diagnosis | Retrospective, hospital medical records |
| *Roth, 1989* |  | Mild <20 mins | Mild: 13-15 |  |  |  |  |  | Prospective, interview on admission to rehab |
| *Sharma, 2014* | Present | Present | <13 | Positive (CT, MRI) | IQ testing |  |  | Require 2 of PTA, LOC and GCS of 13 or below OR positive neuroimaging. OR change in pre-morbid IQ | Prospective neurological examinations<br>Retrospective, hospital medical record review. |
| *Kaminski, 2017* |  |  |  |  |  |  |  |  | Retrospective, unknown |
| *Hagen, 2010* |  | Mild < 5mins<br>Mod:5-24h | Mild: 14-15<br>Mod: 9-14<br>Severe: 3-8 |  | Mild: No focal neurological deficits<br>Mod: Focal neurological deficits |  |  | Require one of GCS <15 or LOC for TBI | Retrospective, hospital medical records |
| *Dionne, 2025* | Mild <24h<br>Mod:1-6d<br>Sev: >7d | Present | Mild:13-15<br>Mod:9-12<br>Sev:3-8 |  |  |  |  | Only one of PTA, LOC or +ve neuroimaging required | Prospective, neurosurgeon evaluation on admission to hospital |
| *Pagni, 1991* |  |  |  |  |  |  |  |  | Retrospective, hospital medical records |
| *Snell, 2023* | Mild <24h<br>Mod:1-6d<br>Sev: >7d |  | Mild: 13-15<br>Mod:9-12<br>Sev:3-8 | CT or MRI* |  |  |  | Neuroimaging only used to separate mild (negative) from mild complicated, mod-sev (present) | Retrospective, hospital medical, emergency medical services, surgical and acute rehabilitation records. |
| *Richards, 1988* |  | Present |  |  |  |  |  |  | Prospective, interview rehab admission |
| *Macciocchi, 2008* | Mild <24h<br>Mod:1-6d<br>Sev: >7d |  | Mild: 13-15<br>Mod:9-12<br>Sev:3-8 | CT or MRI* |  |  |  | Either PTA or +ve neuroimaging required | Retrospective, hospital medical, emergency medical services, surgical and acute rehabilitation records. |
| *Sikka, 2019* | Present | Present | Mild:13-14<br>Mod:9-12<br>Sev:3-8 | Positive | Cognitive deficits | ICD-9 (not specified) |  | One of PTA, LOC or +ve neuroimaging required | Retrospective, hospital medical records |
| *Bombardier 2016* | Present | Mild <30min,<br>Mod:30min-24h,<br>Sev: >24h |  |  | Mental status change |  |  | Improbable TBI = no LOC/mental status change<br>Possible TBI = mentals status, no LOC<br>TBI = LOC | Prospective, interview at rehab admission, (*Step 2 of the TBI-4 Self report*) |
| *Tolonen, 2009* | Mild <24h<br>Mod 1-7d<br>Sev >7d | Mild <30 mins<br>Mod: 30 mins= 6h<br>Severe > 6h | Mild: 13-15<br>Mod:9-12<br>Sev:3-8 | Mild: None<br>Mod: +ve<br>Sev +ve, operated | Mild: No symptoms<br>Mod: Transient<br>Sev: Permanent deficit |  |  | Only one of the criteria needed for diagnosis | Prospective, interview at rehab admission<br>Retrospective hospital record review |
| *Case-matched studies* |  |  |  |  |  |  |  |  |  |
| *Baguley, 2019* |  | Present | <15 | Positive | Positive |  |  | Only one of the criteria needed for diagnosis | Retrospective, hospital admission records |
| *Bradbury, 2008* | Present | Present |  | +ve CT (acute care)<br>+ve MRI# (2-6m) | Positive |  |  | Required +ve neuroimaging for diagnosis | Retrospective, hospital admission records |
| *Macciocchi, 2004* | Present | Present | Mild:13-15<br>Mod:9-12<br>Severe:3-8 | CT imaging |  |  |  | Either LOC or PTA required | Retrospective, hospital admission records |
| *Macciocchi, 2012* | Mild <24h<br>Mod:1-6d<br>Sev: >7d |  | Mild: 13-15<br>Mod:9-12<br>Sev:3-8 | CT or MRI* |  |  |  | Either PTA or +ve neuroimaging required | Retrospective, hospital admission records |
| *Macciocchi 2013* | Mild: PTA <24h |  |  | CT negative |  |  |  | Either PTA or GCS<15 required | Retrospective, hospital medical, emergency medical services, surgical and acute rehabilitation records. |

The information used to diagnose a TBI also varied widely between studies, with 5 studies not specifying the criteria used at all^49–53^. In a further 6 studies TBI was diagnosed using only ICD billing codes^32,54–58^, with 3 of these studies not specifying the codes used^32,55,57^. An additional 2 studies utilised ICD billing codes alongside examination of medical records for terms associated with brain injury like head trauma^59^ or for complications associated with TBI including contusions^60^. In other studies TBI was diagnosed via the presence of post-traumatic amnesia (PTA)^33,34,38,40,41,43,45–47,47,48,61–65^, loss of consciousness^9,10,10,34,38,43,45–48,60,61,63,64,66–68^, Glasgow Coma Score (GCS)^9,33,34,38,40–43,46–48,61,62,64–70^, the presence of focal neurological deficits^38,46,47,63,65–68,71^, cognitive changes^47,48,64,65,72^ or positive neuroimaging^33,34,40,41,44,46–48,61–66,68^. How this information was used to diagnose TBI varied, with two studies requiring the presence of two indicators of head injury^34,48^, while in the majority of studies (n=20) only one indicator of TBI was required for a diagnosis to be made. In most studies this required the presence of one criterion from a predetermined list, typically encompassing one of PTA, an indicator of loss of consciousness or abnormal neuroimaging^33,34,38,41,43,45,46,48,61,64–68^. However, in 8 studies only one criterion was assessed, most commonly a decreased GCS^10,42,69,70^, but studies also diagnosed TBI based solely on abnormalities on neuroimaging^44^, a clinical presentation suggestive of TBI^71^ ^3,15,29–32,36^ and the presence of cognitive deficits^72^.

Sixteen studies then investigated the severity of the TBI associated with an SCI^9,33,38–43,46,47,61,62,64,65,67,70^. In five studies this was based only on length of loss of consciousness/GCS ^9,42,61,64,70^, but more commonly studies incorporated other measures including length of PTA^33,39–41,43,62,65^, neuroimaging findings^33,40,41,46,47,61,62,65^ and severity of neurological and cognitive deficits^38,46,65,67^ to differentiate TBI severity. However, given that many studies only required one indicator of TBI for a diagnosis to be made, severity may have also been based solely off this information^38,39,46,61,62,67^.

### 3.3 Incidence/prevalence

Having identified DDx, 34 studies provided information regarding the incidence of DDx, with incidence ranging from 7.4% in a US hospital-based population^32^ and 74.2% in a Finnish study which specifically examined individuals for indication of prior TBI on admission to rehabilitation and then retrospectively analysed medical records^46^ (Table 2). Overall, twelve studies reported an incidence of DDx of less than 30%^32,38,42,49,54,55,57,59,65,69–71^, seventeen studies between 30-50%^9,10,39,41,43–45,48,50–53,56,58,60,66,67^ and three >50%^33,46,64^. The reported incidence ranges were broadly similar across studies, regardless of methodological quality.

**Table 2:**
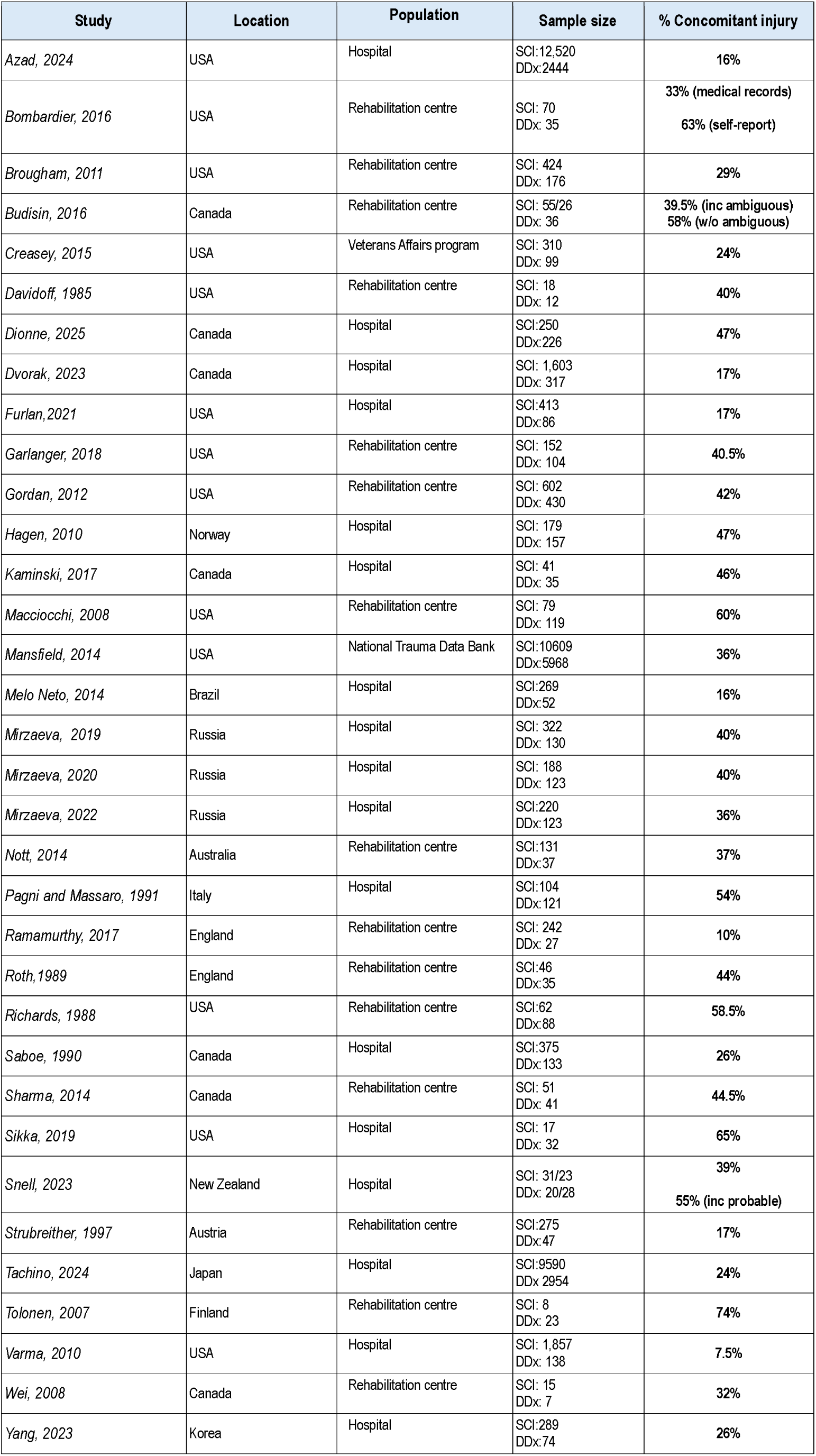
Reported incidence of concomitant TBI with SCI.

High-quality studies reported incidences ranging from 16^42,54^–60%^33^, moderate-quality studies from 7.5^32^–74%^46^, and low-quality studies from 10^38^–65%^64^. Rather, two studies demonstrated that the incidence of DDx depended on the criteria used, Bombardier *et al* reported an incidence of 33% using retrospective review of medical records, but 62% with self-report via the step 2 TBI-4 Self Report interview^47^, although they reported that the interview lacked specificity. Budisin *et* al had a different approach, examining the effect of including or excluding ambiguous cases. Definitive positive TBI required two of the presence of PTA< a period of loss of consciousness, a GCS at admission of 14 or below or positive neuroimaging, whilst ambiguous cases only had one indicator, with this changing the incidence from 39.5 vs 58%^34^.

Indeed, how the TBI was identified may influence the prevalence reported in these studies. The reports where DDx was reported as less than 30% tended to rely on less information to identify the presence of a TBI, including ICD codes in five studies^32,54,55,57,59^, only GCS in three studies^42,69,70^, non-specified symptoms and signs in one study^71^ with another not specifying how TBI was classified at all^49^. Only two studies with an estimated incidence of <30% examined multiple criteria including post-traumatic amnesia, Glasgow Coma Score and clinical presentation^38,65^. In contrast those studies reporting an incidence of 30-50% tended to use more information, with half utilising more than one criteria to assess whether a head injury had occurred^34,39,41,43,45,47,60,66,67^, although four of the studies, including two from the same research group, did not specify how a head injury was classified^50–53^. The studies that identified an incidence of >50% all used multiple criteria including reference to post-traumatic amnesia^33,46,47,64^, decreased Glasgow Coma Score or presence of loss of consciousness^33,46,47,64^, neuroimaging^33,46,64^ and TBI related symptoms^46,47,64^, with two specifically examining hospital medical, emergency medical services, surgical and acute rehabilitation records to increase the likelihood of identifying information suggesting the presence of head injury^33,46^.

Prevalence of DDx may also have been influenced by the cohort sizes examined. The studies with the highest incidence also had low cohort sizes ranging from 31-198^33,46,47,64^, whilst those with the lowest incidence included the studies with the largest cohorts including two papers with 10000-15000 participants^54,70^ and two papers with 1000-5000 participants^32,55^, alongside a further three with a cohort size of 500-1000^49,57,69^

#### 3.3.1 Characteristics of DDx compared to SCI alone

Alongside examining the incidence of DDx, investigation of whether injury characteristics differed between SCI alone and DDx were also examined (Table 3). In the 13 papers differentiating age and/or sex at injury in those with a DDx compared to SCI alone, none reported a meaningful difference in age at injury^33,34,39,41,43–45,54,58,59,64,67,69^. There were mixed reports on whether DDx differentially affected one sex. Five papers reported a similar proportion of males affected by SCI alone compared to DDx ^43,54,58,59,69^, three papers slightly more females in the DDx group, with an increase in the %females within the DDx cohort of 10-24%^34,44,64^ and four slightly more males ^39–41,67^. Two of these papers found that this was severity dependent-with more males in the severe DDx group in the Hagen *et al* paper^67^, and more males in the mild DDx group in the Garlanger et al paper^39^.

**Table 3:**
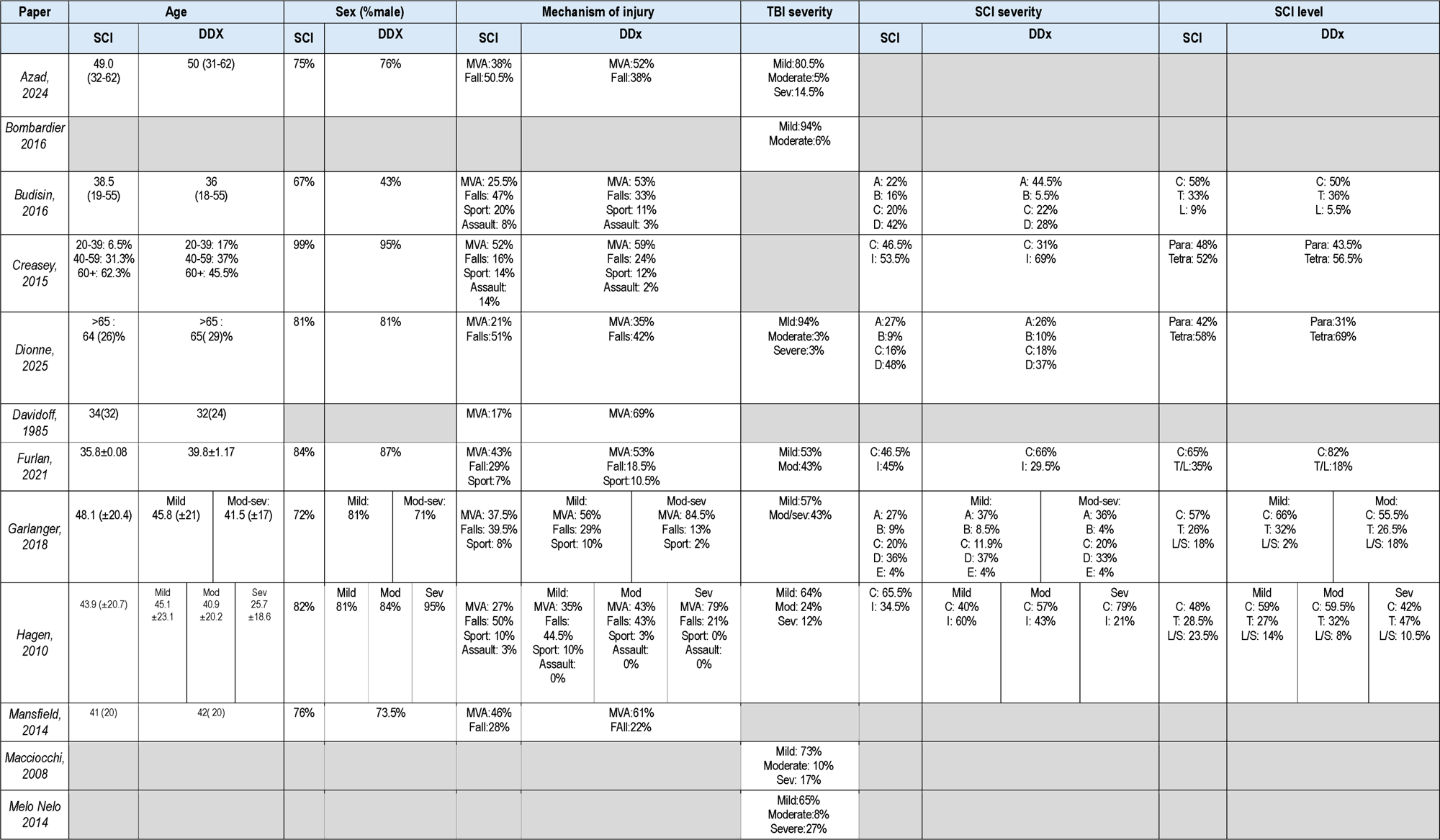
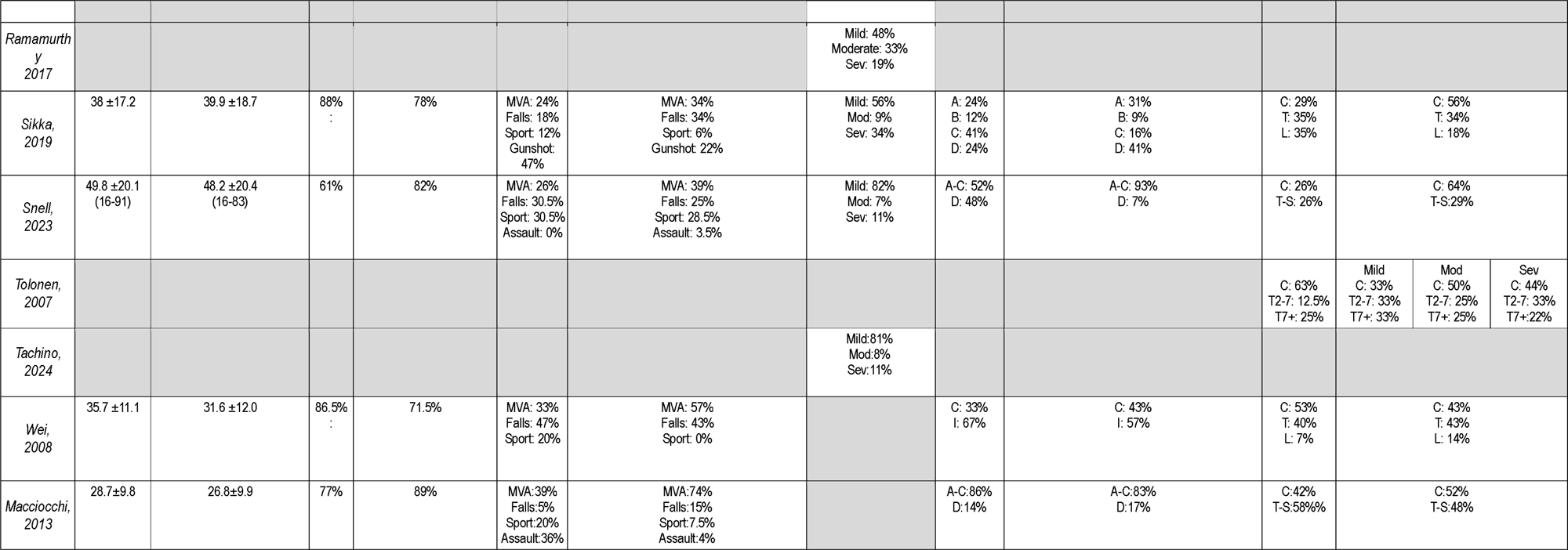
Injury characteristics of individuals with an SCI compared to a DDx.

**Table 4:**
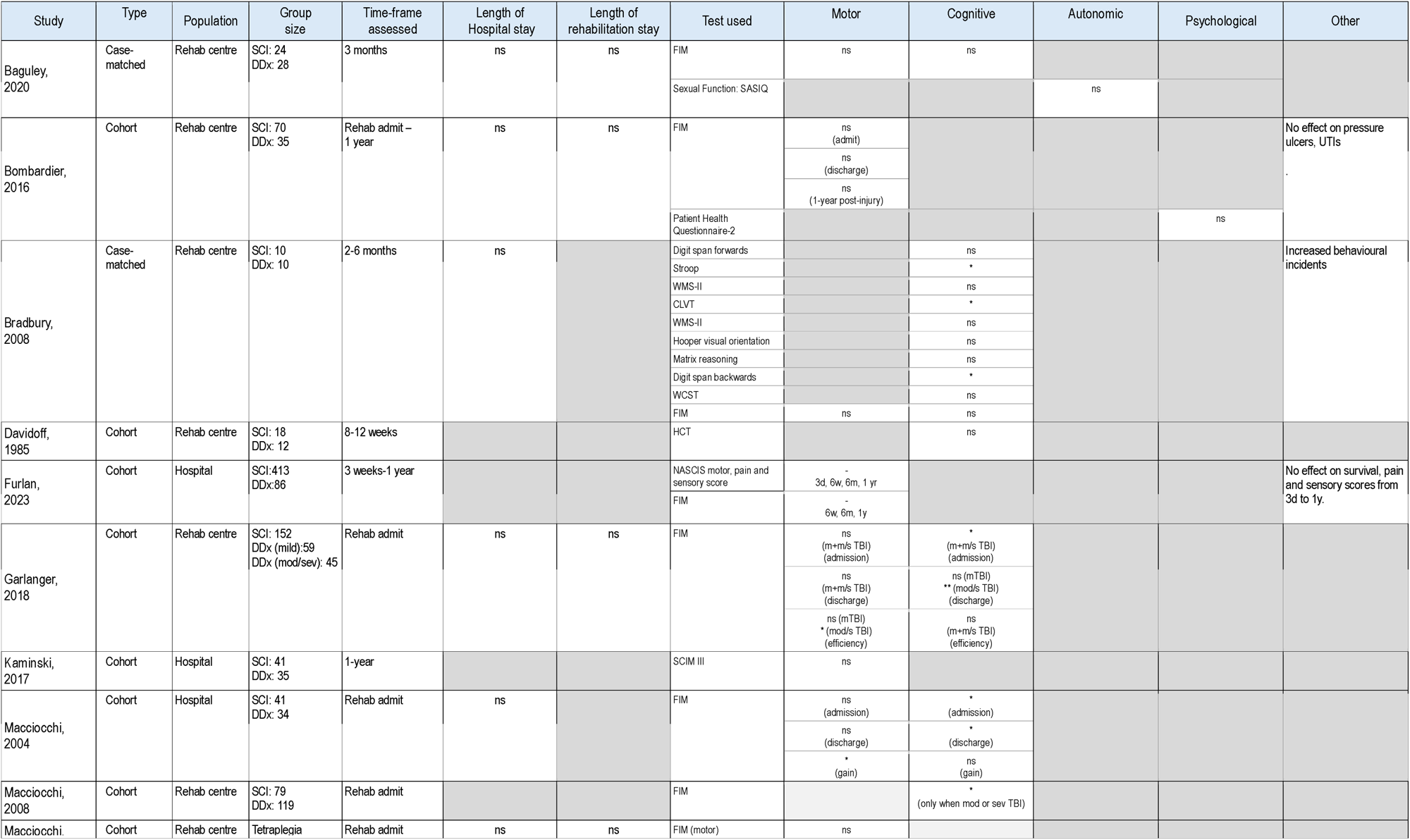
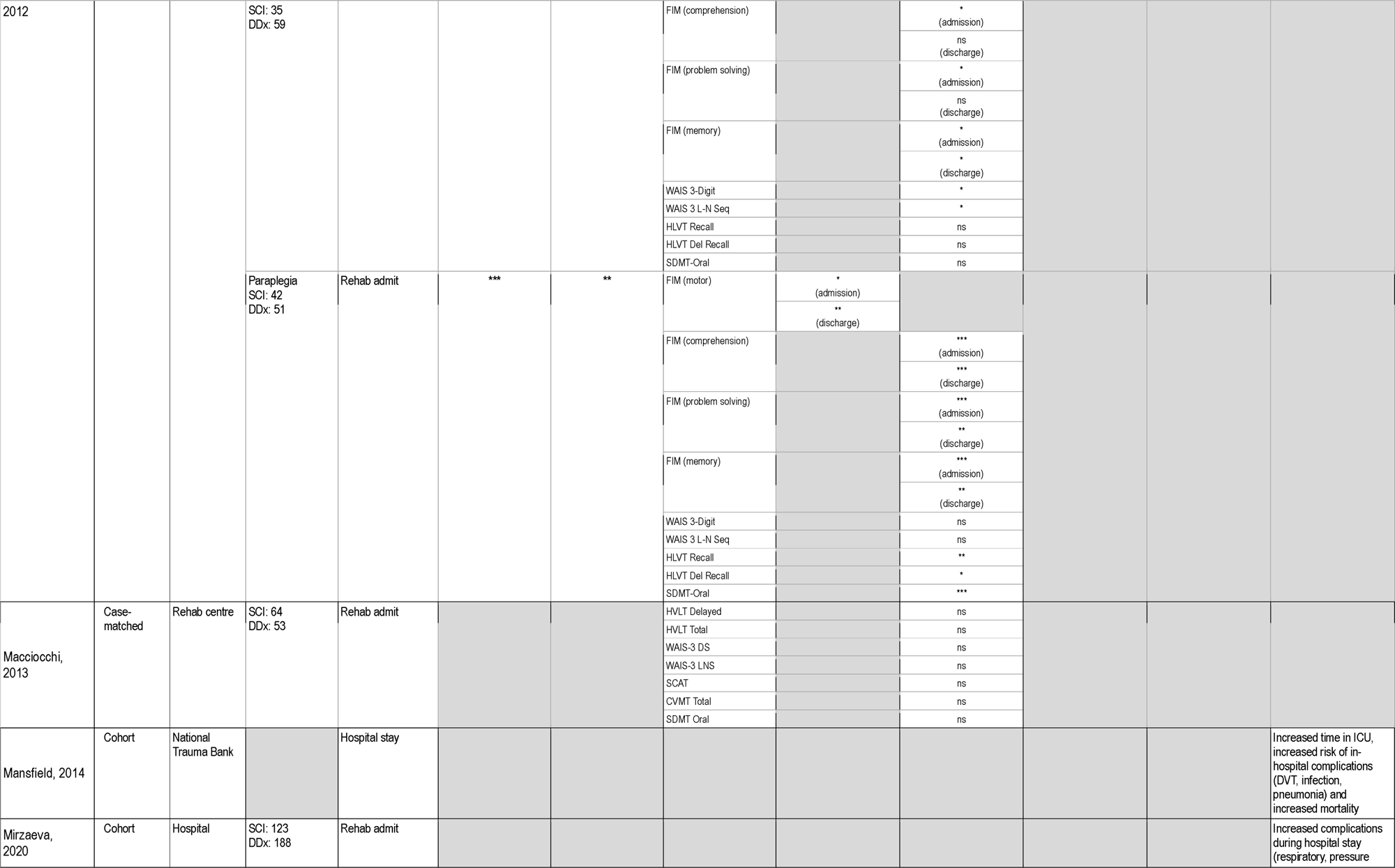
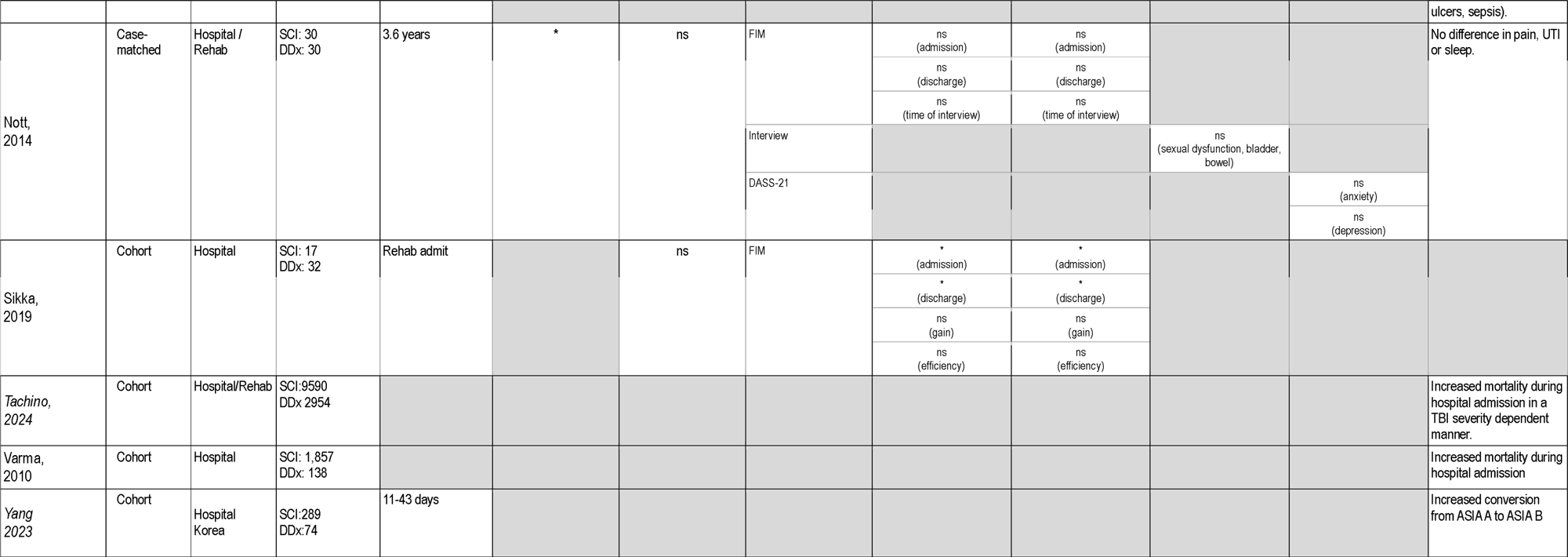
Functional outcomes following a DDx compared to SCI alone.

When comparing DDx to SCI alone, the mechanism of injury differed with DDx linked to higher rates of MVAs compared to SCI alone in 12/13 studies^34,39–41,43–45,54,58,64,67,69^ (34-74% vs 21-52%), with a similar rate of MVA, but slightly more falls in the other study^59^. Where severity of TBI was considered, MVA was more common with increasing severity of TBI^39,67^, increasing from the mechanism of injury in 35-56% of cases in mTBI to 79-84.5% with mod-sevTBI^39,67^.

When looking at severity of TBI in individuals with a DDx mild TBI was the most common severity in 11/13 studies reporting this characteristic^33,38,41–43,47,54,64,67,69,70^, although the reported % of DDx associated with mild TBI varied widely between these from 94% to 48% of those with a DDx. Only one paper reported higher rates of moderate TBI, at 42% of cases compared to 19% mild and 28% severe^65^ and another a relatively even split between the three severities of TBI (26:35:29%). It should be noted in this Tolonen *et al* paper those with a moderate TBI were classified based on length of PTA and LOC, rather than GCS, which was typically still 14-15 and would have been rated as mild according to criteria of other studies^46^.

DDx did not appear to consistently affect the location of the SCI, which was reported as either via anatomical level (cervical, thoracic, lumbar) or as presenting as para or tetraplegia. Four papers reported that those with a DDx had similar ratios of the spinal cord injury location compared to SCI alone^34,39,59,67^, four that DDx was associated with higher rates of cervical injury (52-86 vs 29-65%)^40,41,64,73^ or tetraplegia (69% vs 58%)^43^ and two lower rates of cervical injury in individuals with a DDx ^44,46^, although these studies had small sample sizes of 22-23 participants.

SCI severity was assessed as complete/incomplete in 4 studies and as ASIA score in 7 studies that provided information on DDx compared to SCI alone. Where TBI severity was not considered, rates of complete SCI were reported as slightly higher than DDx in two studies (43-66% vs 33-46.5%)^44,73^ and slightly lower in another study (31 vs 46.5%)^59^ This is potentially TBI severity dependent, with Hagen *et al* finding that with mild or moderate TBI the rates of incomplete SCI were greater than SCI alone, but with severe TBI the rates of complete SCI were greater than SCI alone^67^. Where ASIA scores were reported for each level from A-D, DDx had a slightly higher proportion of individuals scoring A in 3/4 studies^34,39,64^, with this not affected by TBI severity^39^. In two papers A-C scores were grouped together with one paper finding those with an A-C score more commonly had a DDx^41^ and another similar proportions between DDx and SCI alone^33^.

### 3.5 Functional outcomes

18 of the 40 studies included information on whether concomitant injury affected recovery when compared to SCI alone, with 7 assessed as high quality, 4 as moderate quality and 7 as low quality, with small sample sizes, how the TBI was identified and consideration of severity the most common concerns. Measures included within these studies included length of hospital stay and rehabilitation length, alongside more specific measures to evaluate motor, cognitive, autonomic or psychological outcomes.

#### 3.5.1 Hospital and rehabilitation

Acutely, a DDx was found to increase in hospital complications including respiratory infections, pressure ulcers and sepsis^50,58^, with longer stays in intensive care^58^ and an increase in mortality during hospital admission^32,58,70^, although this increase in mortality was lost when individuals with GCS <10 on arrival were excluded^69^. However, despite the increase in complications only 1^66^ of 5^39,47,61,63,68^ studies reported an increase in length of hospital stay, with Nott *et al* finding that those with a DDx spent a mean difference of 21 days longer in hospital than people with an SCI alone. TBI, may also differentially affect SCI depending on the level of injury, with paraplegic, but not tetraplegic individuals with a DDx having a longer length of stay in both hospital and rehabilitation than those with an SCI alone^62^. By the time of rehabilitation admission, however all 8 studies reporting this outcome found no difference in time spent in rehabilitation between DDx and SCI only ^39,47,61–64,66,68^. During rehabilitation, a DDx was associated with a higher rate of spontaneous recovery with more people converting to ASIA A to ASIA B^65^.

#### 3.5.2 Motor and sensory outcomes

10 of the 17 studies investigated motor outcomes^39,45,47,52,61,62,64,66,68,73^, and only one sensory and pain scores^69^. For motor outcomes 8 used the motor portion of the Functional Independence Measure (FIM)^39,47,61–64,66,68,69^, 1 used the Spinal Cord Independence Measure (SCIM III)^52^ and 1 the National Acute Spinal Cord Injury Study motor score by assessing motor function in 14 segments^69^. Sensory function was also assessed by the National Acute Spinal Cord Injury Study test, with sensory function examined via pinprick and light touch sensation for C2-S5, with pain also assessed in six regions^69^.

Motor function assessed by raw FIM scores found no differences at admission to or discharge from rehabilitation in 3 of the 6 studies investigating this time point post-injury^47,61,66^. Of the three studies that reported significantly lower FIM motor scores on rehabilitation admission and discharge, two only reported this effect in a subpopulation of people: paraplegic rather than tetraplegic individuals^62^ and moderate/severe but not mild TBI^39^. Only Sikka *et al* found an overall effect of concomitant injury on raw FIM motor scores regardless of severity of TBI or level of SCI, although notably this paper had one of the highest reported rates of concomitant injury, at 65%^64^.

Effects of concomitant injury on motor outcome during rehabilitation were also presented as FIM gain, the change in score from admission to discharge, or FIM efficiency, this being the change in score considering the length of stay. Macciocchi *et al* found a significant decrease in FIM gain with concomitant injury^61^, although this was not seen in another study that evaluated this measure^64^. Garlanger *et al*, but not Sikka *et al* found a significantly lower FIM efficiency with concomitant injury, although the Garlanger *et al* study only found this in people with a moderate/severe, but not mild TBI suggesting a TBI severity effect^39,64^.

Furlan *et al* examined motor and sensory function using the National Acute Spinal Cord Injury Study tests at 3 days, 6 weeks, 6 months and 1 year and found that although those with a DDx s scored significantly worse than SCI alone at all time-points via Mann-Whitney U-test, this was found to be no longer significant when confounding factors were taken into account via multiple regression analysis^69^. Indeed, four other studies investigated longer time points for motor function, out to 3.5 years post-injury, utilising either the motor FIM score or SCIM III, with no study finding significant change in functional motor outcome with concomitant injury compared to SCI alone, although all studies had overall sample sizes of <100^47,52,63,66^.

#### 3.5.3 Cognitive outcomes

10 studies investigated cognitive outcomes^33,39,40,45,61–64,66,68^ with the most common tool the cognitive portion of the FIM task. Two studies reported significant differences in raw FIM scores at rehabilitation and discharge in all individuals with a DDx^61,64^. However, Nott *et al* found no differences during rehabilitation^66^ and Garlanger *et al*^39^ and Macciocchi *et al* (2008)^33^ found this was severity dependent, with significant differences only in moderate to severe TBI DDx compared to SCI alone. Macciocchi *et al* (2012) examined the specific cognitive subdomains of the FIM task, with all those with a DDx performing significantly worse in comprehension, problem solving and memory at admission compared to SCI alone^62^. However, whilst DDx paraplegic individuals were still significantly different on all domains compared to paraplegic SCI individuals at rehabilitation discharge, people with tetraplegia and head injury were only significantly different in memory^62^.

Two studies utilised a broader array of cognitive assessments during rehabilitation which included the Wechsler Adult Intelligence Scale-3^rd^ edition Digit Span (WAIS-3 DS), WAIS-3 Letter-Number Sequencing (WAIS-3 LNS), Hopkins Verbal Learning Test (HLVT) with total and delayed recall subsections, and the Symbol Digit Modalities Test (SMDT). One study which only included mild TBI DDx found no significant differences^40^ whilst Macciocchi *et al* (2012) found significant differences when level of SCI was taken into account in DDx individuals with varying severity of TBI^62^. DDx associated with tetraplegia led to significantly worse outcomes on the WAIS-DS and WAIS-3 LNS, but not HVLT or SMDT compared to tetraplegia alone, whereas DDX associated with paraplegia led to the opposite with significant differences in the HLVT delayed and total recall and SDMT, but not WAIS-DS or WAIS-3 LNS compared to paraplegia alone^62^.

Four studies investigated time-points beyond rehabilitation, with no significant difference in cognitive FIM scores to 6 months post-injury^63,66,68^ or in the Halstead Category Test at 3 months post-injury^45^. However, a more comprehensive battery assessing broader cognitive domains including attention, verbal and visual memory and executive function, found significant differences in the Stroop test, California Verbal Learning test (CLVT) and Digit Span backwards test at 2-6 months post-injury, but not in Digit-span forwards, the Wechsler Memory Scale-3rd edition (WMS-III), Hooper visual orientation, matrix reasoning or Wisconsin Card Sort Test (WCST) in DDx compared to SCI alone^63^.

#### 3.5.4 Other outcomes

Post-hospital stay DDx did not affect the rate of pressure ulcers^47^, pain^66,69^ or urinary tract infections^66^, nor overall survival^69^, although DDx was associated with an increase in behavioural incidents to six months post-injury in those with a DDx^63^. Two studies also specifically investigated autonomic functions, looking at the effects of DDx from 3 months^68^ to six years post-injury^66^. No significant difference at 3 months was found in sexual function using the Sexuality after Spinal Injury Questionnaire (SASIQ)^68^, with an interview also identifying no significant differences in sexual, bladder, and bowel function up to six years post-injury^66^. Similarly, two studies investigated whether a DDx affected psychological outcomes up to six years post-injury. No significant effects of a DDx compared to SCI alone on depression or anxiety were found utilising the Patient Health Questionnaire-2 at one-year post-injury^47^, or on the DASS-21 questionnaire at 3-6 years post-injury^66^.

## 4. Discussion

This systematic review summarises the clinical evidence to date on the prevalence of concomitant TBI in individuals with traumatic SCI, the barriers to identifying TBI in people presenting with an SCI and its influence on functional outcomes. It is difficult to determine the prevalence of concomitant TBI, as the criteria for assessing TBI differ between studies, and the clinical information required to make a diagnosis are not always collected. Nonetheless, even with these limitations the majority of papers reported an incidence of 30-50% suggesting that concomitant injury is common in the SCI cohort. The effect of concomitant TBI on functional recovery following SCI was also examined, with the articles reviewed here finding little difference in functional recovery, although this should be taken in the context variable quality and low sample sizes of many of the included studies. A moderate-severe TBI was associated with an increase in in-hospital mortality and complications, but overall concomitant injury had minimal effects on recovery trajectory during rehabilitation. Notably few studies have examined the long-term effects of concomitant injury, particularly on cognition, with subtle cognitive deficits detected at six months post-injury in the DDx group, but no later time-points examined. These findings highlight the need for prospectively designed studies to capture the presence of a TBI at the time of SCI, and to follow these individuals chronically, to fully understand the impact of a concomitant TBI on the recovery from SCI.

The prevalence of reported concomitant TBI with SCI varies widely between studies and seems to be most influenced by the criteria used to diagnose the TBI rather than differences in injury characteristics. Of the studies providing details on how diagnosis was made, 80% utilised clinical indices including PTA, loss of consciousness/GCS, neurological symptoms and alterations in neuroimaging. Neuroimaging is most useful in diagnosing moderate-severe TBI, with mTBI only showing abnormalities on neuroimaging in ∼10-20% of cases presenting to the emergency department^3–75^. Indeed, diagnosing mTBI has become increasingly controversial, as there is no universal consensus on a definition. It is generally characterised by a head injury disrupting brain function as evidenced by altered mental status (e.g. confusion and disorientation), loss of consciousness of less than <30 min, PTA of less than anLhour, and/or transient neurological dysfunction (e.g. headache and dizziness PTA^77–79^, but even then LOC may only be found in 8^80^ – 13%^81^ of cases. Beyond this, even with more severe TBI, given that most studies utilised retrospective analysis of medical records there are inherent limitations to being able to identify whether a TBI occurred, including absent or inaccurate documentation. Indeed, two of the studies reporting an incidence of DDx in >50% of people presenting with an SCI, specifically examined hospital medical, emergency medical services, surgical and acute rehabilitation records to increase the likelihood of identifying relevant information^33,41^. Furthermore level of consciousness cannot always be assessed, particularly in people with pre-injury intoxication, reported in ∼30% at time of SCI^82,83^ or those that require sedation and intubation, particularly common in higher cervical SCI injuries^84^. Indeed, only two studies reported assessing for confounds, Tolonen *et al* including intoxication, hypoxia, medication and other medical conditions ^46^, and Macciocchi *et al*, for intubation only^33^. Other retrospective analyses relied only on ICD codes, rather than specific clinical indices, and these typically reported the lowest incidence of concomitant TBI. This may reflect difficulty in capturing the incidence of TBI via this method, with multiple diagnostic codes used to denote TBI, with no consensus on which constitute the best practice^85,86^. Indeed, no research to date has investigated whether care providers in different medical settings use appropriate or consistent codes for TBI^87^. This can be seen by the differing ICD codes included across the studies incorporating this measure. Even in prospective studies identification of mTBI is difficult, with up to 50% of mTBI diagnoses missed in the emergency department setting even in people presenting without an SCI^88–90^. Given the difficulties in capturing the presence of TBI in those presenting with an SCI, and the poor quality of many of the papers included within the study, it is difficult to conclude what the incidence of concomitant TBI is in those presenting with an SCI, highlighting the need to develop consistent methods to accurately diagnose TBI in this cohort.

With the caveat that the studies included herein may thus include individuals with a concomitant TBI in their SCI only cohort, this study also examined whether people with a DDx differed in presentation to people with an SCI only. No differences in age or sex were found, but concomitant TBI was more likely when the mechanism of injury was a motor vehicle accident, accounting for ∼ 80% of moderate severe TBI cases associated with an SCI injury. MVAs generally result in greater impact velocities than other injury mechanisms and lead to head and spinal injuries due to blunt impact and/or inertial loading^91^. Globally, MVAs are the leading cause of both SCI^92,93^ and TBI^94^ Surprisingly unlike previous reports where DDx was associated primarily with cervical injury ^33,49,95,96^, DDx was not consistently associated with a specific injury location in the studies included here, with roughly half supporting an increase in head injury in people presenting with a cervical SCI^40,41,43,64,73^ and half finding either no difference or a decrease in cervical injury^34,39,59,67^. These studies did not consistently differ in geographic location, mechanism of injury or demographic attributes of included participants making it unclear what may drive the difference in injury location between studies. Nonetheless it highlights the need to maintain a clinical suspicion of the potential for a concomitant TBI, regardless of SCI injury location, particularly in high velocity injuries.

Unsurprisingly a concomitant moderate-severe TBI was associated with higher in hospital mortality compared to SCI alone^32,58,70^. This was TBI severity dependent with, Tachino *et al* reporting a mortality rate of 25% in severe TBI (GCS 3-8) compared to 3% in mild TBI (GCS 13-15) DDx cases, with a 10.8% mortality in people presenting with only an SCI^70^. Indeed, when individuals arriving with a GCS < 10 were excluded from analysis no difference in in-hospital mortality was found between those with an SCI only diagnosis and those with a DDx^73^. This supports previous reports suggesting an in-hospital mortality rate for isolated moderate to severe TBI as 15-31%^97,98^ compared to 3-13% for isolated SCI^99–101^. Thus, the increased in-hospital mortality appears to be a consequence purely of the head injury, and not an interaction between the two. The isolated effects of a moderate-severe head injury would also account for the increase in in-hospital complications like pneumonia and sepsis^50,58^ and longer times in ICU. Isolated TBI causes non-neurological complications in approximately 37% of people^102^ compared to ∼24% of those with an SCI only^58^, with respiratory problems the most common^103^. This did not however, lead to significant increases in hospital stay length, or time in rehabilitation in most of the studies. Only Nott *et al* found a significant increase in hospital stay length in all people presenting with a DDx, and this small study had more cervical SCI cases in the DDx group, with higher level of injury itself associated with longer time in hospital^104,105^. This may explain why Macciocchi *et al* found that the effects of concomitant TBI may be more evident in people with paraplegia, with only this subgroup and not those presenting with tetraplegia, having longer length of stay in hospital and rehabilitation.

The effect of concomitant TBI on functional outcomes following SCI were also assessed. Overall concomitant TBI had minimal effects on motor or sensory outcomes either acutely or chronically. Notably motor outcome was assessed with the motor potion of the FIM in 9/10 studies, which assesses activities like eating, grooming, bathing, dressing, toileting, transfers and mobility are scored on a 7-point scale^106,107^. The FIM was developed primarily to assess disability and requirements for burden of care^108^, with ceiling and floor effects for differing lesion levels whereby the scale cannot detect changes in response. For example in paraplegia a ceiling effect is seen in the bed transfer category, whereby no further improvements can be noted, with a floor effect in tetraplegia, where no further deterioration in function can be detected^109^. Indeed overall the FIM has been found to be less sensitive than other tests, in that it is less sensitive to improvement than the quadriplegia index of function ^110^, less able to detect changes in walking than the walking index for spinal cord injury ^111^ and less sensitive to functional changes than the SCIM^112,113^. Nonetheless given that SCI disrupts conduction of sensory and motor signals across the site of the lesion^114^, any additional effects of a disruption of central motor pathways by TBI will be difficult to detect. Indeed, the studies surveyed here suggested that a TBI only worsened acute motor recovery, when the TBI was moderate-severe^39,66^ or in individuals with paraplegia^62^ where disruption of upper motor function caused by the TBI would be more evident. Indeed, motor symptoms are minimal with isolated mild TBI, the most common severity noted with concomitant injury. Following mTBI subtle acute^115,116^ and chronic alterations^117,118^ are found in postural stability, with no effect on gross motor function^119^, which is therefore unlikely to alter FIM motor scores.

Unlike motor function, the cognitive portion of the FIM, evaluating comprehension, expression, social interaction, problem solving and memory on a 7 point scale was able to detect differences between those presenting with a concomitant injury compared to SCI only during rehabilitation, but only in those with moderate-severe TBI, not a mild injury^39,40,61,62,64^. This is not surprising given that TBI is associated with severity dependent cognitive deficits, particularly in attention and executive function^120,121^. Indeed, although mTBI is associated with measurable impairment in various cognitive domains including executive function, learning/memory, attention, processing speed, the deficits are subtle and likely to require more sensitive neuropsychological testing^122^. Even with more severe injury, ceiling effects were observed when the cognitive portion of the FIM was used at later time-points beyond three months post-injury^63,66,68^ with Bradbury *et al* finding that 90% of their cohort had reached the highest score of 35, with no capacity to differentiate between those with an DDx compared to an SCI only. When more sensitive neuropsychological tests were used in this cohort, cognitive performance was worse in people with a DDx compared to SCI alone, with impairments on the Stroop test, CLVT and Digit span backwards^63^, suggesting attention and learning and memory difficulties. To date these tools are yet to be used beyond six months post-injury making it difficult to determine if a TBI at the time of SCI causes long-term cognitive impairments. SCI has been associated with cognitive impairment in domains such as attention, concentration, abstract reasoning, verbal learning, new learning and memory and processing speed in up to 40% of individuals^123^. For example Cohen *et al* found that individuals with an SCI at least one year prior were worse in in fluid composite scores (a measure of executive function), psychomotor speed, and episodic memory^124^, with Craig e*t al* similarly finding that SCI was associated with impaired attention, visoconstructional skills, memory, executive functioning, and language^125^. Although concomitant TBI is commonly proposed as a potential mechanism driving the development of chronic cognitive deficits following SCI^126,127^, as can be seen here, there have been no specific studies to date investigating this topic. Given that SCI alone may also impair cognition, due to factors including autonomic dysfunction driving cerebral blood flow impairment, spread of inflammation to key cognitive areas like the hippocampus^128^, alongside the use of medications including sedatives, anxiolytics, antispasmodics, serotoninergic agents, narcotics, anticonvulsants, tricyclic antidepressants, and skeletal muscle relaxants which are prescribed more often in those with SCIs to the general population^124,129^, this is an important topic that requires further investigation.

Other notable findings here were a differing in the profile of cognitive deficits observed in individuals presenting with tetraplegia compared to paraplegia alongside a TBI at least during rehabilitation, with impaired working memory in DDx associated with tetraplegia and learning and memory in DDx associated with paraplegia compared to SCI alone. This aligns with reports specifically investigating the effects of SCI on cognition at least one year post-injury, with significantly poorer new learning and memory noted in those with paraplegia and impaired processing speed in those with tetraplegia^130^, suggesting that a concomitant TBI may exacerbate or accelerate underlying SCI induced deficits. Why spinal injury level may affect different cognitive domains is not yet understood, higher thoracic/cervical injuries are associated with autonomic dysfunction that can reduce blood pressure^131^ and also impair neurovascular coupling, impairing the matching of blood flow to regional metabolic demands particularly in high demand tasks like cognitive processing^132^. In contrast paraplegia has been suggested to increase cerebral vascular resistance^133^, with the two differing cardiovascular risk factors potentially affecting different brain regions and thus affecting different cognitive domains^134^. Given that TBI also decreases neurovascular coupling^135,136^ and increases cerebral vascular resistance^137,138^, it may act to exacerbate these changes.

Like the paucity of studies investigating chronic effects on cognition, there is a similar lack of studies looking at the effect of concomitant TBI beyond one-year post-SCI on other outcomes, with only Nott *et al* examining out to 3 years post-injury. Nott *et al* found no differences in autonomic dysfunction, anxiety, depression or sleep in a small case-matched sample of individuals with an SCI and those with a DDx^66^, with Bombardier *et al* similarly finding no difference in rates of depression at one-year post-injury^47^. Although these studies used screening tools to assess depression and anxiety, which are not diagnostic, in the Patient Health Questionaire-2^47^ and Depression, Anxiety and Stress Scale^66^, they been shown to be valid in both SCI^139^ and TBI populations^140^ Both TBI and SCI have been associated with increased rates of psychiatric disease. Depression affects approximately 19–26% of individuals living with SCI with this figure approximately three times greater than that reported in the general population^125,141^. Similarly, longitudinal studies show that up to 62% of those who suffer moderate-to-severe TBI are diagnosed with one or more psychiatric illnesses within five years post-injury, of which depression and anxiety disorders are most common^142^. Prevalence of affective disorders is also higher following mTBI relative to population base rates^143^. Indeed, recent meta-analyses have found that individuals with a history of TBI are 1.9 times as likely to experience clinical anxiety^144^, and over twice as likely to experience depression^145^. However here, no synergistic effect between TBI and SCI were found, although larger studies looking at more chronic time-points may be required.

This review of the clinical prevalence of concomitant TBI in people presenting with an SCI identified several barriers to the diagnosis of DDx. Moderate-severe TBI had the greatest impact on in hospital recovery, with increased mortality and complications, with minimal effects of concomitant injury detected during rehabilitation, or to three years post-injury. However given the difficulty of detecting TBI at the time of SCI, these results may be influenced by inclusion of individuals with a concomitant injury in the SCI group. Furthermore, only a small number of studies to date have been conducted, with half of poor quality, highlighting the need for larger studies, over longer time periods to fully understand the implications of the impact of concomitant TBI on the recovery of SCI and how these people can best be supported through their recovery.

## Author Contributions

KS: conceptualisation, methodology, investigation, manuscript preparation-original draft, RG: methodology, investigation, manuscript preparation-review, RO’D: conceptualisation, supervision, manuscript preparation-review and editing; FC: conceptualisation, supervision, manuscript preparation-original draft, manuscript preparation-review and editing AL: conceptualisation, supervision, manuscript preparation-review and editing

## Ethical Approval

N/A

## Data Availability

All data produced in the present study are available upon reasonable request to the authors

