## Supplementary material for "Double Trouble - The prevalence of concomitant traumatic brain injury in individuals with spinal cord injury and its impact on functional outcomes: a systematic review": Supp Table 1 and 2

Table 1: Adaption of the National Institutes of Health (NIH) quality assessment tool for observational cohort and cross-sectional studies used to assess methodological quality

| Major Components | Response options | | | |
| --- | --- | --- | --- | --- |
| 1. Was the research question or objective in this paper clearly stated? | Yes | No | Cannot Determine | Not applicable |
| 2. Was the study population clearly specified and defined? | Yes | No | Cannot Determine | Not applicable |
| 3. Was the participation rate of eligible persons at least 50%? | Yes | No | Cannot Determine | Not applicable |
| 4. Were all the subjects selected or recruited from the same or similar populations (including the same time period)? Were inclusion and exclusion criteria for being in the study prespecified and applied uniformly to all participants? | Yes | No | Cannot Determine | Not applicable |
| 5. Was a sample size justification, power description, or variance and effect estimates provided? | Yes | No | Cannot Determine | Not applicable |
| 6. For the analyses in this paper, were the exposure(s) of interest measured prior to the outcome(s) being measured? | Yes | No | Cannot Determine | Not applicable |
| 7. Was the timeframe sufficient so that one could reasonably expect to see an association between exposure and outcome if it existed? | Yes | No | Cannot Determine | Not applicable |
| 8. For exposures that can vary in amount or level, did the study examine different levels of the exposure as related to the outcome (e.g., categories of exposure, or exposure measured as continuous variable)? | Yes | No | Cannot Determine | Not applicable |
| 9. Were the exposure measures (independent variables) clearly defined, valid, reliable, and implemented consistently across all study participants? | Yes | No | Cannot Determine | Not applicable |
| 10. Was the exposure(s) assessed more than once over time? | Yes | No | Cannot Determine | Not applicable |
| 11. Were the outcome measures (dependent variables) clearly defined, valid, reliable, and implemented consistently across all study participants? | Yes | No | Cannot Determine | Not applicable |
| 12. Were the outcome assessors blinded to the exposure status of participants? | Yes | No | Cannot Determine | Not applicable |
| 13. Was loss to follow-up after baseline 20% or less? | Yes | No | Cannot Determine | Not applicable |
| 14. Were key potential confounding variables measured and adjusted statistically for their impact on the relationship between exposure(s) and outcome(s)? | Yes | No | Cannot Determine | Not applicable |

Table 2: Assessment of quality for each study indicating whether they exhibited the attribute (green, did not (red), it could not be assessed (yellow) or was not applicable (grey).

|  | Country | Type of study | Research Objective | Study population | Participation Rate | Subjects | Sample size | Timeframe | Different levels of severity | Independent variables | Outcome measures | Blinding | Attrition Rate | Statistical analysis |
| --- | --- | --- | --- | --- | --- | --- | --- | --- | --- | --- | --- | --- | --- | --- |
| Azed, 2024 | US | CS |  |  |  |  |  |  |  |  |  |  |  |  |
| Baguely, 2019 | Aus | CS |  |  |  |  |  |  |  |  |  |  |  |  |
| Bombardier, 2016 | US | CS |  |  |  |  |  |  |  |  |  |  |  |  |
| Bradbury, 2008 | Canada | C |  |  |  |  |  |  |  |  |  |  |  |  |
| Brougham, 2011 | US | CS |  |  |  |  |  |  |  |  |  |  |  |  |
| Budisin, 2016 | Canada | CS |  |  |  |  |  |  |  |  |  |  |  |  |
| Creasey, 2015 | US | CS |  |  |  |  |  |  |  |  |  |  |  |  |
| Davidoff, 1985 | US | Coh |  |  |  |  |  |  |  |  |  |  |  |  |
| De Melo Neto, 2014 | Braz | CS |  |  |  |  |  |  |  |  |  |  |  |  |
| Dionne, 2025 | Can | CS |  |  |  |  |  |  |  |  |  |  |  |  |
| Dvorak, 2023 | Can | Coh |  |  |  |  |  |  |  |  |  |  |  |  |
| Furlan, 2021 | US | Coh |  |  |  |  |  |  |  |  |  |  |  |  |
| Garlanger, 2018 | US | CS |  |  |  |  |  |  |  |  |  |  |  |  |
| Gordan, 2012 | US | CS |  |  |  |  |  |  |  |  |  |  |  |  |
| Hagen, 2010 | Norway | Coh |  |  |  |  |  |  |  |  |  |  |  |  |
| Kaminski, 2017 | US | CC |  |  |  |  |  |  |  |  |  |  |  |  |
| Mansfield, 2014 | US | Coh |  |  |  |  |  |  |  |  |  |  |  |  |
| Macciocchi, 2004 | US | Coh |  |  |  |  |  |  |  |  |  |  |  |  |
| Macciocchi, 2008 | US | Coh |  |  |  |  |  |  |  |  |  |  |  |  |
| Macciocchi, 2012 | US | Coh |  |  |  |  |  |  |  |  |  |  |  |  |
| Macciocchi, 2013 | US | Coh |  |  |  |  |  |  |  |  |  |  |  |  |
| Melo Nelo, 2014 | Brazil | Coh |  |  |  |  |  |  |  |  |  |  |  |  |
| Mirzaeva, 2019 | Russia | Coh |  |  |  |  |  |  |  |  |  |  |  |  |
| Mirzaeva, 2020 | Russia | Coh |  |  |  |  |  |  |  |  |  |  |  |  |
| Mirzaeva, 2022 | Russia | Coh |  |  |  |  |  |  |  |  |  |  |  |  |
| Nott, 2014 | Austria | CC |  |  |  |  |  |  |  |  |  |  |  |  |
| Pagini 1991 | Italy | CS |  |  |  |  |  |  |  |  |  |  |  |  |
| Ramamurthy, 2017 | UK | CS |  |  |  |  |  |  |  |  |  |  |  |  |
| Richards, 1988 | US | Coh |  |  |  |  |  |  |  |  |  |  |  |  |
| Roth, 1989 | US | Coh |  |  |  |  |  |  |  |  |  |  |  |  |
| Saboe, 1990 | Can | CS |  |  |  |  |  |  |  |  |  |  |  |  |
| Sharma 2014 | Can | CS |  |  |  |  |  |  |  |  |  |  |  |  |
| Sikka, 2019 | US | CS |  |  |  |  |  |  |  |  |  |  |  |  |
| Snell, 2023 | NZ | Coh |  |  |  |  |  |  |  |  |  |  |  |  |
| Strubreither, 1997 | Austria | CS |  |  |  |  |  |  |  |  |  |  |  |  |
| Tachino, 2024 | Japan | CS |  |  |  |  |  |  |  |  |  |  |  |  |
| Tolonen, 2007 | Finland | CS |  |  |  |  |  |  |  |  |  |  |  |  |
| Varma, 2010 | US | CS |  |  |  |  |  |  |  |  |  |  |  |  |
| Wei, 2008 | Canada | CC |  |  |  |  |  |  |  |  |  |  |  |  |
| Yang, 2023 | Korea | CS |  |  |  |  |  |  |  |  |  |  |  |  |
